# Radiomic and clinical predictors of cachexia in non-small cell lung cancer patients treated with immunotherapy

**DOI:** 10.1101/2020.10.08.20207415

**Authors:** Wei Mu, Evangelia Katsoulakis, Kenneth L. Gage, Chris J. Whelan, Matthew B. Schabath, Robert J. Gillies

## Abstract

**Background:** Cachexia is present in up to 50% of patients with cancer and may contribute to primary resistance to immunotherapy. Biomarkers to predict cachexia are urgently required for early intervention. Herein, we test the hypothesis that pre-treatment ^18^F-FDG-PET/CT-based radiomics can be used to predict cachexia and subsequently associated with clinical outcomes among patients with advanced non-small cell lung cancer (NSCLC) who are treated with immunotherapy.

**Methods:** This retrospective multi-institution study included 210 patients with histologically confirmed stage IIIB-IV NSCLC who were treated with immune checkpoint blockade between June 2011 and August 2019. Baseline (pre-immunotherapy) PET/CT images of 175 patients from Moffitt Cancer Center were used to train (N=123) and test (N=52) a radiomics signature to predict cachexia, which was also used to predict durable clinical benefit (DCB), progression-free survival (PFS) and overall survival (OS) subsequently. An external cohort that enrolled 35 patients from James A. Haley Veterans’ Hospital (VA) was used to further validate the predictive and prognostic value of this signature.

**Results:** A radiomics signature demonstrated cachexia prediction ability with areas under receiver operating characteristics curves (AUC) of 0.77 (95%CI:0.68-0.85), 0.75 (95%CI:0.60-0.86) and of 0.73 (95%CI:0.53-0.92) in the training, test and external VA cohorts, respectively. For the further investigation of prognostic value, this signature could identify the patients with DCB with AUC of 0.67 (95%CI:0.57-0.77), 0.66 (95%CI:0.51-0.81), and 0.72 (95%CI:0.54-0.89) in these three cohorts. Additionally, the PFS and OS were significantly shorter among patients with higher radiomics signature in all the three cohorts (p<0.05).

**Conclusion:** Using PET/CT radiomics analysis, cachexia could be predicted before the start of the immunotherapy, making it possible to monitor the patients with a higher risk of cachexia and identify patients most likely to benefit from immunotherapy.

## Introduction

Cachexia, a syndrome that induces progressive functional impairment(1), is present in up to 50% patients with cancer(2), and accounts for 20% of cancer-related deaths(3). Cachexia is defined as more than 5% weight loss over 6 months, or more than 2% weight loss if body mass index (BMI) is less than 20 kg/m^2^ in absence of simple starvation(4). Compared to other solid neoplasms, such as breast and thyroid cancer, and haematological malignancies, the prevalence of cachexia is higher in patients with lung cancer(5, 6), which is one of the most prevalent cancer types and a leading cause of cancer-related death(7). Cancer cachexia not only increases patients’ mortality, but also impairs the response to first and second line chemo-/radiotherapy(5). Specifically, the presence of cachexia in patients treated with immune checkpoints blockade agents (ICIs), which has significant improved the long term survival for the patients with advanced non-small cell lung cancer (NSCLC) (8-10), may promote primary resistance to immunotherapy because of suboptimal drug exposure due to the hypothesis that the anorexia/cachexia-related metabolic wasting may accelerate antibody blood clearance (11, 12). Therefore, early identification of patients likely to develop cachexia is critically needed by the scientific community to initiate the interventions as early as possible to optimize the treatment of cachexia and attenuate its development (13). Early identification of cachexia-prone patients may also help predict prognosis of ICIs to guide treatment(5).

To identify patients with high risk of developing to cachexia at early stage, prior studies have attempted to define criteria for pre-cachexia, a stage when early clinical and metabolic signs such as anorexia and inflammation were present, but substantial weight loss was not (4), utilizing weight loss, body mass index (BMI)(14), anorexia, systemic inflammation(15), biochemistry, food intake, activities and functional status (16). However, these studies were analyzed based on overall survival, and couldn’t be used for cachexia prediction.

The main causes of cancer-associated cachexia are systemic metabolic modifications including excess catabolism, increased energy expenditure and inflammation, which are determined by tumor-driven molecular alterations (11, 17). Additionally, metabolic alterations associated with malignant disease may alter lymphocyte function by limiting the availability of key nutrients(18). As a metabolic imaging technique, ^18^F-FDG PET/CT is sensitive in reflecting metabolic changes. 18F-FDG uptake assessed by PET/CT has been shown to be also associated with cellular and molecular alterations(19), and to reflect not only the number of lymphocytes, but also the activation state of the lymphocytes themselves(20). Further, the recent “radiomics” analysis, which converts medical images into high-dimensional mineable data(21), can successfully predict tumor gene mutations (22-25) and protein expression (26). However, the association between PET/CT based radiomics and cachexia hasn’t been well investigated. Therefore, we hypothesized that radiomics analysis of the primary tumor from pre-treatment PET/CT images can predict the probability of developing cachexia.

In this study, we utilize NSCLC patients receiving ICIs, some of whom developed cachexic during their therapeutic course, were utilized to develop a radiomics model to predict cachexia during the course of immunotherapy and to investigate the prognostic value of the generated radiomics signature.

## Patients and Methods

### Patients

Retrospective patient cohorts were identified from two institutions: H. Lee Moffitt Cancer Center & Research Institute (HLM) and the James A. Haley Veterans’ Hospital (VA), both in Tampa, Florida. Inclusion criteria included patients with histologically confirmed advanced stage (IIIB and IV) NSCLC who were treated with anti-PD1 or anti-PD-L1 immune checkpoint blockade between June 2011 and August 2019. The detailed exclusion criteria are provided in **Figure 1** and include: (a) PET/CT images not available before the start of immunotherapy; (b) ≥ 3 months between the PET/CT acquisition and the start of immunotherapy; (c) other treatments that were performed between imaging acquisition and start of treatment; (d) follow-up time of <6 months after start of treatment; (d) no weight record six month after the start of the immunotherapy. Thus, 175 patients from HLM, who was split into training (N=123) and test cohorts (N=52) randomly by 70-30% and 35 external patients (test) from the VA were used to train and test the radiomics signatures to predict cachexia, and to investigate the association of the radiomics signature and the clinical outcomes.

**Figure 1.**
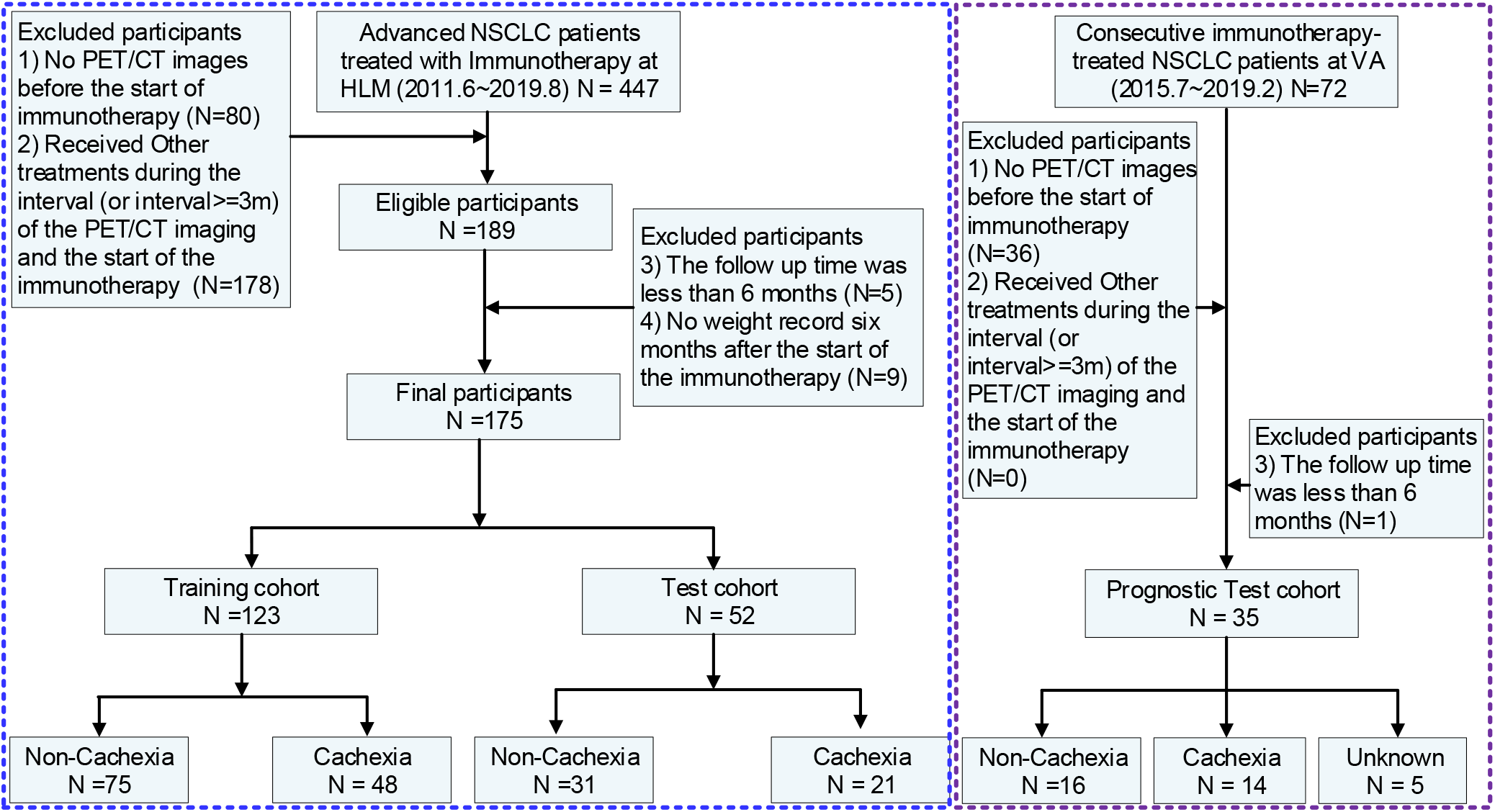
Diagram of study participant enrollment.

Clinical characteristics including age at diagnosis, sex, body mass index (BMI), smoking status, history of COPD (chronic obstructive pulmonary disease), Eastern Clinical Oncology Group (ECOG) performance status (PS), and sites of distant metastasis (M) were obtained from the medical records. BMI was categorized according to: 20.0 kg/m^2^, underweight; 20.0 to 24.9 kg/m^2^, normal weight; 25.0 to 29.9 kg/m^2^, overweight; and 30.0 kg/m^2^, obese(27). Cachexia was defined as patients with more than 5% weight loss over past 6 months, or more than 2% weight loss with body mass index (BMI) less than 20 kg/m^2^. Progression free survival (PFS) and overall survival (OS) defined as the time from the start date of immunotherapy to progression (defined according to Response Evaluation Criteria in Solid Tumors (RECIST1.1)) and death, were chosen as the end points of the study. Patients free of progression (or death) or lost to follow-up were censored at the time of last confirmed contact.

This study was approved by the Institutional Review Boards at University of South Florida (USF) and the James A. Haley Veterans Hospital, and was conducted in accordance with ethical standards of the 1964 Helsinki Declaration and its later amendments. The requirement for informed consent was waived, as no PHI is reported.

### PET/CT Image Analysis

The details of PET/CT imaging analysis are presented in supplementary section 1 and the pipeline of this study is provided in **Figure 2**. Briefly, all PET images were converted into SUV units by normalizing the activity concentration to the dosage of ^18^F-FDG injected and the patient body weight after decay correction. Muscles were identified in two to four adjacent axial images within the CT series at the third lumbar vertebra, including rectus abdominus, abdominal (lateral and oblique), psoas, and paraspinal (quadratus lumborum, erector spinae) by a 5-year experienced orthopedist (Y. W.). These images were refined with a Hounsfield unit (HU) range of −29 to 150 Human averaged for each patient. The total muscle cross-sectional area (cm^2^) was normalized for height in meters squared (m^2^) and reported as lumbar Skeletal Muscle Index (SMI) in cm^2^/m^2^ (27).

**Figure 2.**
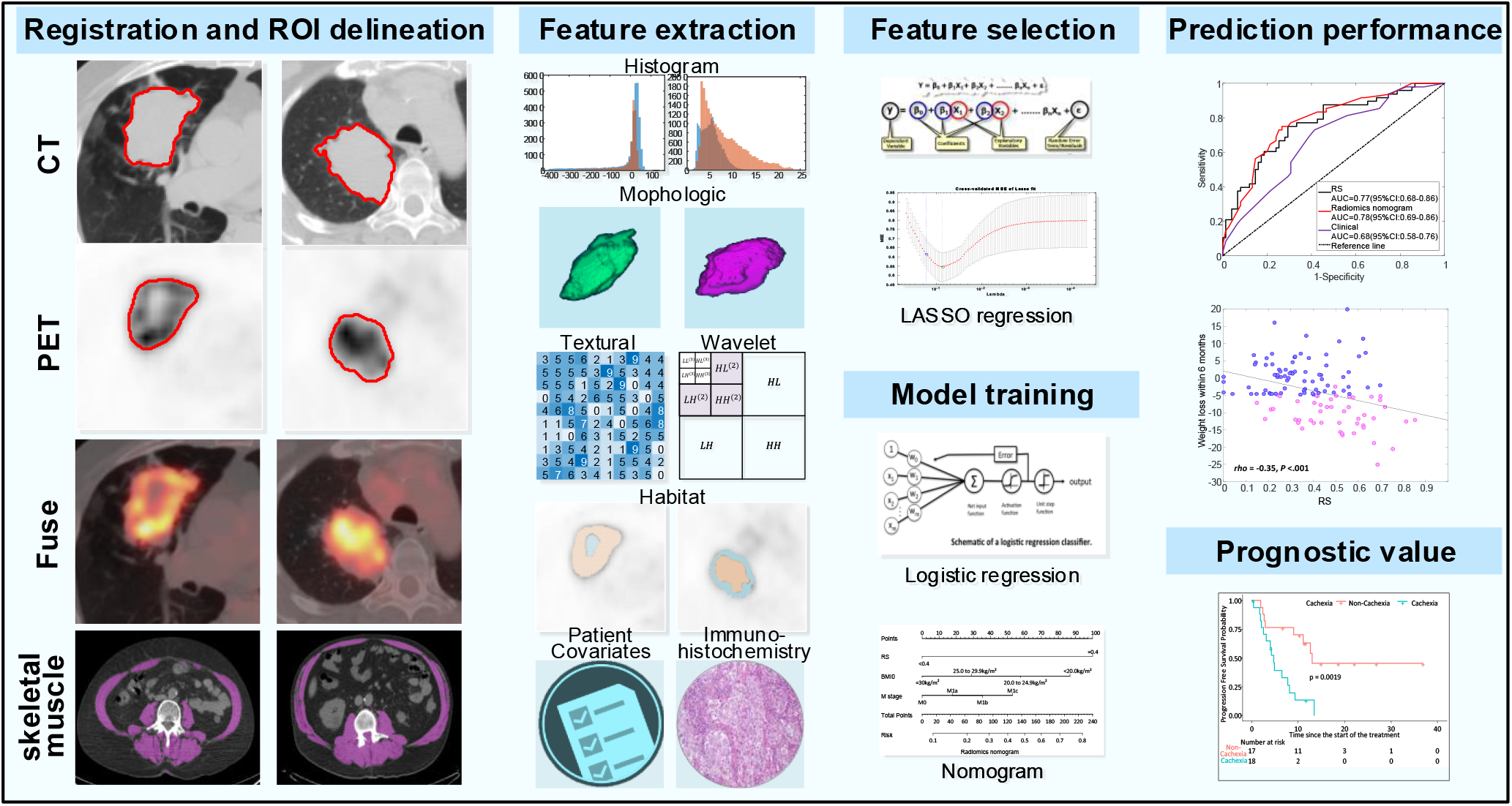
Radiomics Workflow. The workflow includes registration and automatic delineation, imaging preprocessing and feature extraction, feature selection, model training and model validation.

The primary lung tumors of PET and CT images were semi-automatically segmented with an improved level-set method based on the gradient fields (28), and 30 randomly selected nodules in the training cohort were segmented twice. After spatial registration using a rigid transformation by maximizing the Dice Similarity Coefficients on the condition that the maximal axial cross sections of the tumor nodules were aligned, fusion images were generated from the fused PET and CT images on a voxel-wise basis (29). Additionally, on each tumor-restricted PET image, Otsu thresholding (30) was performed to automatically maximize inter-class variance. Using the obtained threshold, the corresponding tumor of PET images was divided into high and low metabolic (SUV) regions, representing distinct habitats. The masks of these two habitats were then mapped to the co-registered CT images, and two CT sub-regions were subsequently obtained. Therefore, four sub-regions regions of PET and CT images including PEThigh, PETlow, CThigh, and CTlow were included. Consequently, 1053 quantitative features (31) (364 whole-tumor PET features, 364 whole-tumor CT features, 65 fusion features, and 260 habitat imaging based features) were extracted. Details are provided in Supplementary Section 2.

### Feature Selection and Construction of the Radiomics Signature

To reduce the dimension of the radiomics features, a four-step feature selection procedure was performed to obtain the key radiomic features to predict cachexia. Firstly, the inter-rater agreement of the radiomics features were calculated by intraclass correlation coefficient (ICC) between segmentations, and only the features with ICC larger than 0.8 were retained. Then, a two-sample T test was used to pre-select the radiomics features that were significantly (P<0.05) different between the cachexia and non-cachexia patients. Next, to reduce redundancy, the significantly different were grouped according to the absolute value Pearson correlation coefficient (denoted by |r|) of 0.9, and only the single feature with the largest classification ability, based on area under the receiver operating characteristics curve (AUC), in each group were selected to be representative (the “Avatar”) for that group Finally, a Least Absolute Shrinkage and Selection Operators procedure (LASSO)-logistic regression analysis was used to select the most useful predictive features with non-zero coefficients, and generate the radiomics signature (RS) through a linear combination weighted by the corresponding coefficients (32). The penalty parameter (λ) in LASSO was selected using 10-fold cross validation by minimum mean cross-validated error.

### Statistical Analysis

The Wilcoxon signed-rank test and Fisher’s exact test were used to test the differences for continuous variables and categorical variables, respectively. Pearson correlation coefficient was used to test for correlations between continuous variables. The cutoffs of RS and SMI used for the classification were determined by maximizing Youden’s index based on the training cohort. Univariable and multivariable logistical regression models (backward step-down selection with Akaike information criterion as the stopping rule)(33), which were presented as radiomics nomograms, were used to assess the predictive ability of RS, SMI, clinical common used metrics including SUVmax, MTV (metabolic tumor volume) and volume (from CT images), and other clinical characteristics (categorized BMI, sex, age, smoke status, COPD, ECOG, distant metastasis, and histology). The area under the receiver operating characteristics curve (AUC), accuracy, sensitivity, specificity, and the 95% confidence interval (CI) by the Delong method(34) were used to assess the ability of different models in discriminating between cachexia and non-cachexia patients. To demonstrate the significantly incremental value of different models, total net reclassification improvement (NRI) was calculated. For PFS and OS comparison, Kaplan-Meier analysis and log-rank test were used. P-value less than 0.05 was regarded as significant, and statistical analyses were conducted with R (version 3.5.1) and MATLAB (R2019b).

## Results

### Clinical characteristics

The clinical characteristics of the patients used to train and test the predictor for cachexia are presented in **Table 1**. Among these 175 patients (96 males, 79 females), the mean age was 66 ± 12 years with a median PFS and OS of 8.40 and 27.6 months, respectively. Of these, 69 patients experienced more than 5% weight loss (weight change: −10.28 ± 4.63 kg/m^2^) during the first six months of immunotherapy, and were regarded as cachectic. All 35 of the external VA patients (**Supplementary Table S1**) are male, with the mean age of 71.40 ± 7.19 years. Ten patients had stage IIIB disease, while the remaining 25 patients were stage IV. The median PFS and OS are 8.13 months and 13.10 months, respectively. Of the 29 patients who had their weight recorded six months after the start of the immunotherapy, 14 of them suffered from cachexia with an accompanying weight loss of 8.93 ± 3.16 kg/m^2^.

**Table 1.** Demographic and clinical characteristics of patients.

| Characteristic | Training Cohort |  | p<sup>†</sup> | Test Cohort |  | p<sup>†</sup> | p<sup>‡</sup> |
| --- | --- | --- | --- | --- | --- | --- | --- |
|  | Cachexia<br>(N=48) | No-Cachexia<br>(N=75) |  | Cachexia<br>(N=21) | No-Cachexia<br>(N=31) |  |  |
| Age(y) |  |  | 0.21 |  |  | 0.33 | 0.62 |
| Mean ± SD | 64.40±11.31 | 67.35±9.71 |  | 62.33±17.04 | 66.06±15.15 |  |  |
| Sex, NO. (%) |  |  | 0.85 |  |  | 1.00 | 0.41 |
| Male | 28 (58.33) | 42 (56) |  | 10 (47.62) | 16 (51.61) |  |  |
| Female | 20 (41.67) | 33 (44) |  | 11 (52.38) | 15 (48.39) |  |  |
| BMI (kg/m<sup>2</sup>) |  |  | **0.016\*** |  |  | 0.30 | 0.62 |
| Mean ± SD | 25.1±5.55 | 26.97±4.43 |  | 25.53±3.47 | 26.41±5.89 |  |  |
| BMI category (kg/m<sup>2</sup>) |  |  | **0.018\*** |  |  | .066 | 0.81 |
| <20.0 | 6 (12.5) | 3 (4) |  | 2 (9.52) | 0 (0) |  |  |
| 20.0 to 24.9 | 21 (43.75) | 25 (33.33) |  | 7 (33.33) | 14 (45.16) |  |  |
| 25.0 to 29.9 | 15 (31.25) | 29 (38.67) |  | 10 (47.62) | 10 (32.26) |  |  |
| ≥30 | 6 (12.5) | 18 (24) |  | 2 (9.52) | 7 (22.58) |  |  |
| Skeletal muscle index (SMI) (cm<sup>2</sup>/m<sup>2</sup>) |  |  | 0.46 |  |  | 0.17 | 0.50 |
| Mean ± SD | 41.52±14.61 | 42.57±13.35 |  | 37.65±11.98 | 44.74±16.79 |  |  |
| ECOG PS |  |  | **0.05\*** |  |  | 0.18 | 0.83 |
| 0 | 6 (12.5) | 23 (30.67) |  | 2 (9.52) | 8 (25.81) |  |  |
| 1 | 40 (83.33) | 51 (68) |  | 18 (85.71) | 23 (74.19) |  |  |
| ≥2 | 2 (4.17) | 1 (1.33) |  | 1 (4.76) | 0 (0) |  |  |
| Distant Metastasis |  |  | **0.017\*** |  |  | 0.64 | 0.45 |
| M0 | 9 (18.75) | 25 (33.33) |  | 4 (19.05) | 9 (29.03) |  |  |
| M1a | 6 (12.5) | 18 (24) |  | 3 (14.29) | 7 (22.58) |  |  |
| M1b | 21 (43.75) | 20 (26.67) |  | 11 (52.38) | 12 (38.71) |  |  |
| M1c | 12 (25) | 12 (16) |  | 3 (14.29) | 3 (9.68) |  |  |
| Histology, NO. (%) |  |  | 0.077 |  |  | 0.58 | 0.09 |
| ADC | 27 (56.25) | 55 (73.33) |  | 12 (57.14) | 15 (48.39) |  |  |
| SCC | 21 (43.75) | 20 (26.67) |  | 9 (42.86) | 16 (51.61) |  |  |
| Weight change within 6 months |  |  | <.001* |  |  | <.001* | 0.96 |
| Mean ± SD | -10.54±4.65 | 0.86±5.21 |  | -9.69±4.63 | 0.5±3.98 |  |  |
| Smoke, NO. (%) |  |  | 0.84 |  |  | 0.87 | 0.16 |
| Non-smoker | 18 (37.5) | 31 (41.33) |  | 5 (23.81) | 9 (29.03) |  |  |
| Former smoker | 28 (58.33) | 42 (56) |  | 14 (66.67) | 20 (64.52) |  |  |
| Current smoker | 2 (4.17) | 2 (2.67) |  | 2 (9.52) | 2 (6.45) |  |  |
| COPD |  |  | 0.14 |  |  | 0.72 | 0.66 |
| NO. (%) | 11 (22.92) | 9 (12) |  | 3 (14.29) | 7 (22.58) |  |  |
| Best Response |  |  | **0.05\*** |  |  | **< 0.001\*** | 0.67 |
| PR/CR/SD | 13 (27.08) | 9 (12) |  | 10 (47.62) | 1 (3.23) |  |  |
| PD | 35 (72.92) | 66 (88) |  | 11 (52.38) | 30 (96.77) |  |  |
| Progression-free Survival |  |  | **0.001\*** |  |  | **0.005\*** | 0.84 |
| Median (IQR) | 5.37<br>(2.60-13.77) | 11.93<br>(7.43-NR) |  | 3<br>(1.77-9.63) | 16<br>(6.17-50.20) |  |  |
| Overall Survival |  |  | **0.011\*** |  |  | **0.009\*** | 0.30 |
| Mean(95%CI) | 30.99<br>(5.18-20.84) | 38.75<br>(31.12-46.37) |  | 24.19<br>(11.90-36.47) | 38.72<br>(28.03-49.40) |  |  |
|  | Cachexia<br>(N=48) | No-Cachexia<br>(N=75) |  | Cachexia<br>(N=21) | No-Cachexia<br>(N=31) |  |  |
| RS |  |  | <.001* |  |  | 0.003* | 0.77 |
| Median (IQR) | 0.51 (0.40, 0.62) | 0.31(0.22,0.43) |  | 0.49(0.39,0.60) | 0.32(0.22,0.43) |  |  |

Note.- : RT is short for radiotherapy; Chemo is short for chemotherapy; SD: standard deviation; IQR is short for interquartile range; ADC is short for adenocarcinoma and SCC is short for squamous cell carcinoma; NR means not reached. Data are patient numbers, with percentages in parentheses. The comparasion of continours variables between two groups was performed with Wikcoxon sign rank test, and the categorical variables were compared with Fisher's test. The P value for PFS and OS are derived from the log-rank test; P<sup>†</sup> value is derived between Cachexia and non-Cachexia; P<sup>‡</sup> is derived training and test cohorts. Data are patient numbers, with percentages in parentheses; \* means P value <.05; The demographic and clinical characteristics of external VA patients were provided in Supplemental Table S1.

### Feature Selection and radiomics signature development

Twenty-seven non-redundant features were found to be significantly different between the cachexic and non-cachexic patients in the training set and these were entered into the LASSO analysis. Of these, nine features were selected out of training to construct the radiomics signature, and these were incorporated into the calculation formula (supplementary section 4). Representative images and radiomics signatures (RS) of two patients from baseline PET/CT scan are shown in **Figure 3A-B**.

**Figure 3.**
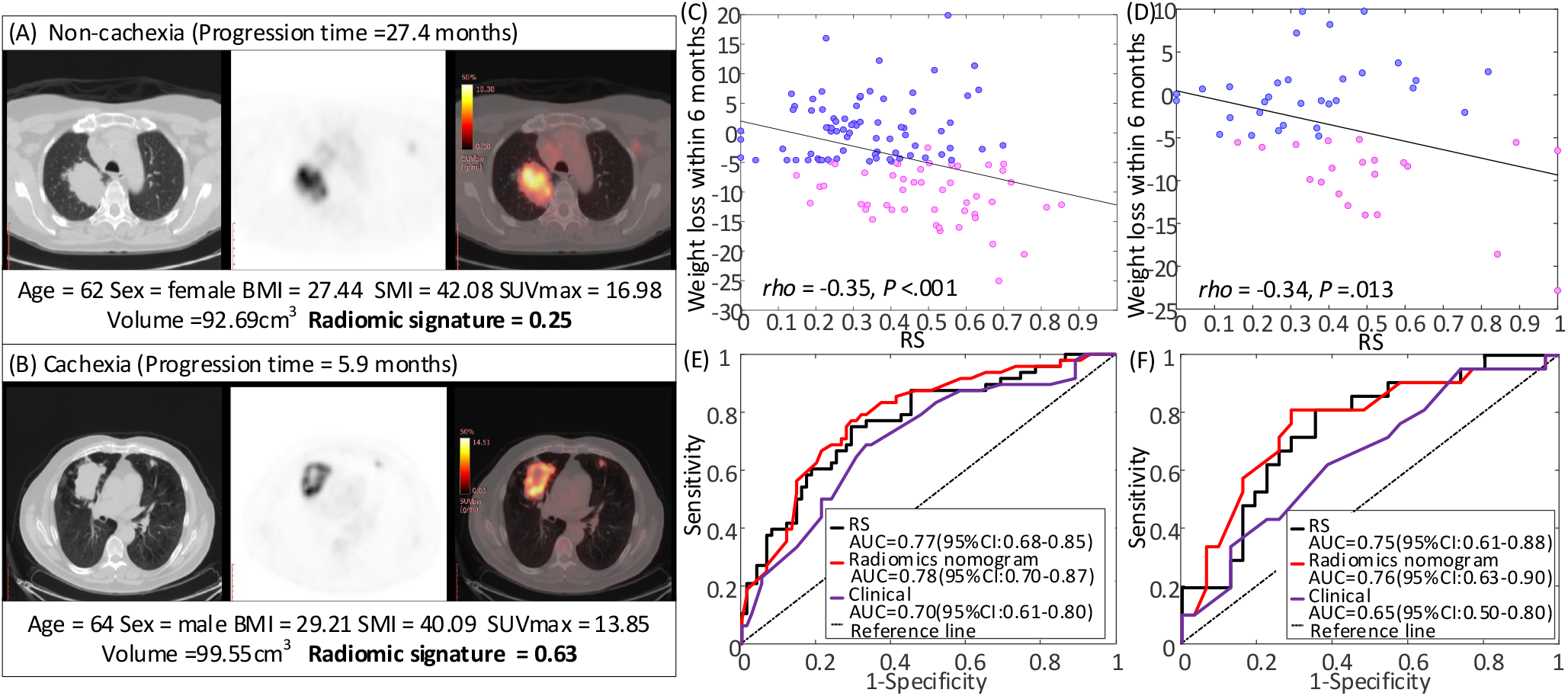
Radiomics signatures of NSCLC patients and their diagnostic performance of in various cohorts. (A) and (B) are the CT, PET, and fusion images for a patient with NSCLC, together with the corresponding clinical variables and radiomics signatures. (C) and (D) are the Spearman’s rank correlation between RS and the weight loss within 6 months since the start of the immunotherapy in the training and test cohorts, respectively. P value indicates two-sided Spearman rank-correlation test. (E) and (F) are the ROC curves of RS, radiomics nomogram and clinical models in the training and test cohorts, respectively.

### Diagnostic performance of the radiomics signature

The RS was significantly different between cachexic and non-cachexic patients in the training cohort (p<0.001), which was validated in the test cohort (p=0.003) and external test cohort (p=0.040). Though the RS was obtained based on the binarized cachexia vs. non-cachexia, the RS was also found to be weakly but significantly correlated with the actual weight change within 6 months since the start of the immunotherapy in both training (Pearson rho = −0.35, p<.001) and test cohorts (Pearson rho = −0.34, p=.013) as shown in **Figure 3C-D**, indicating that a larger RS corresponds to a larger weight loss. This RS yielded an AUC of 0.77 (95%CI: 0.70-0.84) and 0.75 (95%CI: 0.64-0.85) in the training and test cohorts, respectively, and an accuracy of 72.36% (95%CI: 64.23-80.49) and 71.15% (95%CI: 59.62-82.69) in the training and test cohorts, respectively. Detailed information of radiomics signature performance is shown in **Table 2**, and the corresponding ROC curves are shown in **Figure 3E-F**. Additionally, this signature also had a good performance in the external VA cohort with an AUC of 0.73 (95%CI: 0.53-0.92), accuracy of 72.41% (95%CI: 55.17-89.29%), sensitivity of 76.92% (95%CI:53.85-100%), and specificity of 68.75% (95%CI:43.75-87.50%).

**Table 2.** Performance of different models in cachexia prediction.

|  | AUC (95% CI) | ACC (95% CI) | SEN (95% CI) | SPEC (95% CI) | AIC |
| --- | --- | --- | --- | --- | --- |
| Radiomics signature |  |  |  |  |  |
| training | 0.77 (0.68-0.85) | 72.36 (64.23-80.49) | 75.00 (62.5-87.50) | 70.67 (60.00-81.33) | 140.36 |
| test | 0.75 (0.6-0.86) | 71.15 (59.62-82.69) | 80.95 (61.9-95.24) | 64.52 (48.39-80.65) | 65.29 |
| Radiomics Nomogram |  |  |  |  |  |
| training | 0.78 (0.69-0.86) | 73.17 (64.23-80.49) | 77.08 (64.58-87.50) | 70.67 (60.00-80.63) | 142.92 |
| test | 0.76 (0.61-0.89) | 75.00 (61.54-86.54) | 80.95 (61.9-95.24) | 70.97 (54.84-87.1) | 62.78 |
| Clinical nomogram |  |  |  |  |  |
| training | 0.70 (0.62-0.79) | 67.48 (59.35-75.61) | 68.75 (56.25-81.25) | 66.67 (56.00-77.33) | 156.94 |
| test | 0.65 (0.49-0.8) | 61.54 (48.08-75.00) | 61.9 (42.86-80.95) | 61.29 (45.16-77.42) | 69.83 |

Though quantitative indices (BMI and SMI) have been significantly associated with cachexia in other studies(27), neither was significant in our study. BMI achieved an AUC of 0.63 (95%CI: 0.52-0.73, p=0.016) and 0.50 (95%CI: 0.34-0.66, p=0.98) in the training and test cohorts, respectively. SMI provided no significant discrimination ability with AUC of 0.54 (95%CI: 0.43-0.65, p=0.46) and 0.61 (95%CI: 0.46-0.77, p=0.17) in the training and test cohorts, respectively.

Among the HLM patients, 73 patients had weight record of 6 months before the start of the immunotherapy, and 16 of them had history of cachexia. Therefore, a stratified analysis only focusing on the patients without history of cachexia (n=57) was performed. For these patients, the RS, BMI, and SMI achieved different AUCs of 0.76 (95%CI:0.60-0.92, p=0.009), 0.77 (95%CI:0.60-0.94, p=0.006), and 0.82 (98%CI: 0.68-0.95, p=0.001), but the differences among them were not statistically different (RS vs SMI: p=0.57, RS vs BMI: p=0.89, BMI vs SMI: p=0.74, Delong test).

### Clinical prediction model and decision curve analyses

For the univariable logistical regression analysis of the clinical variables, categorized BMI, ECOG PS, and distant metastasis were identified as strong predictors for cachexia (p=0.018, p=0.017 and p=0.015, respectively, **Table S2**). A clinical prediction model was thus trained by incorporating these three clinical variables using multivariable logistic regression analysis which achieved AUC of 0.70 (95%CI: 0.62-0.79) and 0.65 (95%CI: 0.49-0.80) in the training and test cohorts, respectively (Details are shown in **Table 2**).

We then incorporated the RS into the clinical prediction model using further multivariate logistic regression analysis (**Supplementary Table S2**), and this is presented as radiomics nomogram shown in **Figure 4A**. The combined model significantly improved the prediction ability with AUCs of 0.78 (95%CI: 0.69-0.86, p=0.03, Delong test) and 0.76 (95%CI: 0.61-0.89, p=0.047, Delong test) in the training and test cohorts, respectively (Details are shown in **Table 2**). Further, the inclusion of RS yielded a total net reclassification improvement (NRI) of 0.91 (95%CI: 0.59-1.23, p<0.001) and 0.91 (95%CI: 0.43-1.39, p<0.001) in the training and test cohorts, respectively, which further showed significantly improved classification accuracy for cachexia prediction over clinical variables alone. Qualitatively, calibration curves (**Figure 4C)** indicate the agreement between the estimated probability and the actual cachexia rate based on the training (p=0.37) and test (p=0.99) cohorts, respectively.

**Figure 4.**
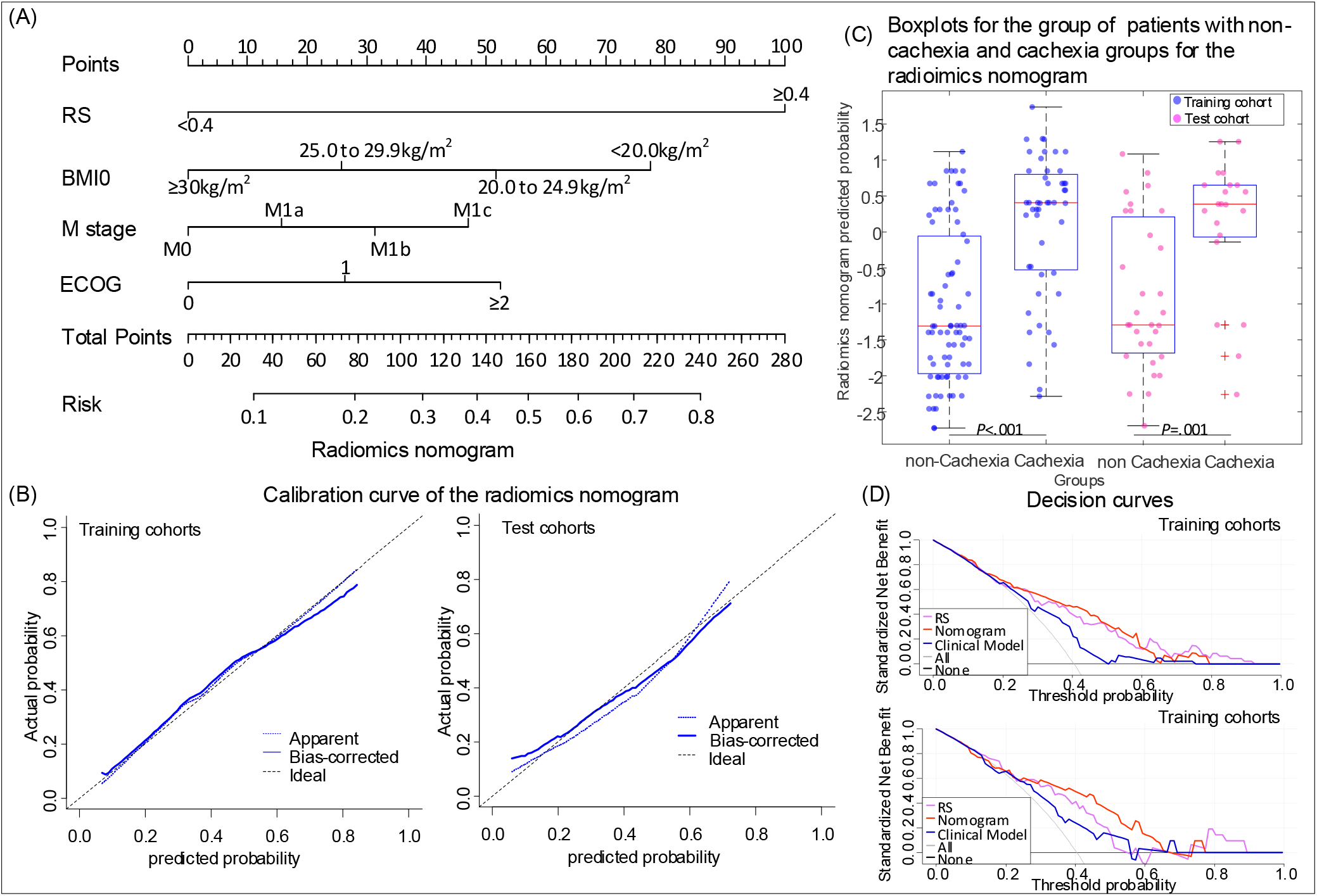
Radiomics Nomograms. The nomogram constructed with RS and clinical variables to estimate the risk of cachexia(A), along with the assessment of the model calibration in the augmented training cohort as well as the training and test cohorts (B). (C) is the box plot of nomogram predicted cachexia probability of individual. In the box plots, the central line represents the median, the bounds of box the first and third quartiles, and the whiskers are the interquartile range. The blue points represent the training cohort, and the magenta points represent the test cohort. P value shows two-sided Wilcoxon signed-rank test. (D) is the decision curves of RS, radiomics nomogram and clinical model in the training and test cohorts.

Decision curve analyses (**Figure 4D**) show the performance of the RS, clinical model, and radiomics nomogram model in clinical application in both training and test cohorts. These show that the radiomics nomogram model showed significant advantages than schema wherein all patients or no patients are assumed to have cachexia. When comparing the three models, the radiomics nomogram model had the highest overall net clinical benefit across the threshold probabilities within the range of 0.30-0.60 in both cohorts.

However, there were no significant differences in predictive ability in both training (p=0.58) and test cohorts (p=0.79) when comparing the RS and the radiomics nomogram. Even for the 57 patients without history of cachexia, the radiomics nomogram achieved higher but also not significant AUC of 0.86 (95%CI: 0.74-0.97, p=0.081, Delong test), compared to the RS alone. Therefore, only RS was used for the following prognostic investigation.

### Prognostic value of the radiomics signature in immunotherapy

The RSs of the patients who experienced durable clinical benefit (DCB, PFS>6 months) were significantly lower compared to those who did not in both the training (0.32 vs. 0.46, p<0.001) and test (0.37 vs. 0.43, p=0.060) cohorts, similar results could be found in the external VA test patients (0.38 vs 0.45, p=0.056). The AUCs of the RS to identify the DCB patients were 0.67 (95%CI: 0.57-0.77, p=0.002), 0.66 (95%CI: 0.51-0.81, p=0.0045), and 0.72 (95%CI: 0.54-0.89, p=0.0029) in the training, test and external VA test cohorts, respectively.

For the training patients, the PFS and OS were significantly longer among patients with low RS (<0.40) versus patients with high RS (PFS: hazard ratio [HR]:1.73, 95%CI: 1.10-2.73, p=0.018; OS: HR: 2.32, 95%CI:1.17-4.63, p=0.017). Among patients with low RS, the median PFS was 11.00 months compared to 5.90 months or patients with high RS (PFS: P=0.016, **Figure 5A**). Median OS was not reached in the low RS group and was 19.77 months in the high RS group (p=0.014, **Figure 5B**). For the test patients, similar results could also be observed in training cohort with HR of 2.18 (95%CI: 1.09-4.38, p=0.028) and 2.62 (95%CI: 1.05-6.56, p=0.040) for PFS and OS estimation, respectively. Median PFS of the patients with low RS was significantly longer with 17.00 months versus 4.17 months (p=0.024, **Figure 5C**), and the median OS was also not reached in the low RS group and was 19.77 months in the high RS group (p=0.014, **Figure 5D**). Through log-rank test, there was not significant difference between RS and real occurrence of cachexia. The external VA test patients further validate the prognostic value of RS with HRs of 2.63 (95%CI: 1.02-6.83, p=0.046) and 3.84 (95%CI: 1.56-9.43, p=0.049, **Figure 5E**) for PFS (4.77 vs 12.97 months, p=0.002, **Figure 5F**) and OS (8.37 vs 22.17 months, p=0.039), respectively.

**Figure 5.**
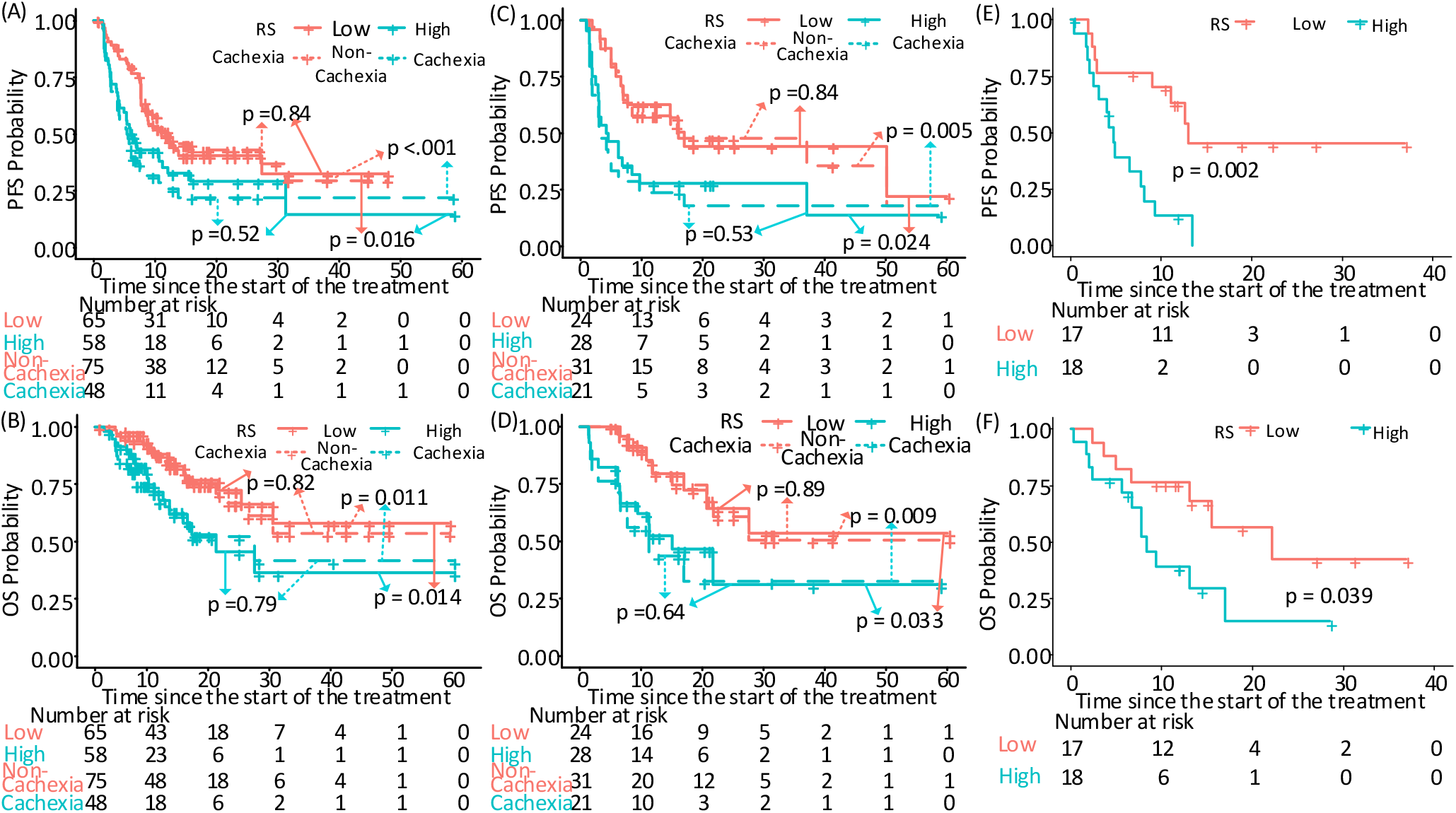
Prognostic ability of RS in various cohorts. (A) and (B) The Kaplan-Meier survival curves of PFS and OS relative to the RS, and cachexia in the training cohort, respectively. (C) and (D) The Kaplan-Meier survival curves of PFS and OS relative to the RS, and cachexia in the test cohort, respectively. (E) and (F) The Kaplan-Meier survival curves of PFS and OS relative to the RS in the external VA cohort, respectively. Comparisons of the above PFS and OS curves were performed with a two-sided log-rank test.

## Discussion

In this study, we found that BMI, distant metastasis, ECOG, and the radiomics signature (RS) of diagnostic PET/CT images, were significant and independent cachexia predictors in patients with advanced stage NSCLC treated with immunotherapy. Importantly, these data can be routinely captured during patient workup as standard of care. The radiomics signature developed in this work further showed significant prognostic value. Therefore, this approach has the potential to be used to optimize patient management and treatment plan with early interventions prior to the start of immunotherapy. As far as we know, this is the first radiomics study of cachexia prediction.

Cancer therapy, could limit tumor growth to suppress the tumor-associated processes responsible for weight loss, but can also induce weight loss due to the potential interference with pathways of muscle anabolism, which means it is difficult to separate cancer cachexia from the effects and complications after cancer therapy. Though not directly a component of cancer-associated cachexia, treatment-associated weight loss is best viewed as an integral part of the syndrome (1). Therefore, the weight loss since the start of the immunotherapy was used to define the cachexia of the enrolled NSCLC patients treated with ICIs. Though cachexia is a multifactorial syndrome defined by an ongoing loss of skeletal muscle mass, the causes of cachexia are usually determined by tumor-driven molecular alterations (11, 17) and tumoral factors have been shown to be important in cancer cachexia(35). Therefore, we hypothesize that the primary tumor could reflect most of the genetic and micro-environmental information. Given that radiomics is a promising approach in characterizing tumors, it is reasonable to develop a radiomics signature to predict cachexia using radiomics analysis of the primary nodule.

Given cachexia is defined primarily by unintended weight loss, several studies have investigated and shown the cachexia-indicative value of initial weight loss (15, 36-38). Additionally, BMI(36), ECOG PS, biochemistry (high C-reactive protein, leukocytes, hypoalbuminemia, or anemia) (37), cancer type, and COPD(38) have also been used to construct cachexia prediction models. Further, CT-based SMI was shown to be predictive of cachexia and survival of patients with cancer(27). However, none of these variables characterized the metabolic changes, characteristic of cancer-associated cachexia. As far as we know, this is the first study to predict cachexia using baseline PET/CT images before the start of the immunotherapy.

When investigating the relationship between clinical characteristics and cachexia, BMI, ECOG PS, and distant metastasis were highly correlated with cachexia, which are consistent with most other studies. However, in contrast to other studies, SMI showed no statistically significant difference between cachexia and non-cachexia (27). A further stratified sub-analysis of patients without a prior history of cachexia showed that SMI was a significant predictor (OR: 0.76, 95%CI: 0.62-0.93, p=0.008) and could achieve high AUC in predicting cachexia. This may indicate that SMI is a good predictor for those patients who hadn’t yet developed cachexia. Additionally, tumor volume and metabolic tumor volume were also not significant predictors, which is consistent with prior observations that cachexia was not tumor size dependent (39, 40). For the investigation of initial weight loss, though the weight loss of the past 6 months and 2 months were also significant factors with OR of 0.85 (95%CI: 0.71-1.01, p=0.056) and 0.78 (95%CI: 0.61-1.00, p=0.049) in the patients who have history weight loss record before the start of the immunotherapy in our study, this factor was not included in the multi-variable analysis in our study due to its unavailability for most of the patients.

The present study does possess some limitations, however. First, because the sample size of patients with history weight loss record was small relative to the entire cohort, which means the predictive value of initial weight loss for the whole cohort couldn’t be incorporated into the prediction model. However, initial weight loss is unavailable for some patients, especially for the patients who received first-line immunotherapy after the diagnosis, which means the clinical use of a model with initial weight loss may be limited. Second, the patient cohorts were heterogeneous in terms of PET/CT image acquisition. However, this also be viewed as a strength of the current approach, and the prognostic value in the external test cohort showed that the developed radiomics signature is robust and transportable.

## Conclusions

In conclusion, an effective radiomics signature from pretreatment PET/CT images has been identified and may serve as a predictive biomarker to identify both patients who have larger risk of developing cachexia after the start of the immunotherapy, and patients most likely to benefit from immunotherapy. Due to the advantage of being based on routinely acquired patient information and its non-invasive characteristic, this signature could be used for optimizing the treatment plan pending in larger and prospective trials.

## Data Availability

The PET/CT imaging data and clinical information are not publicly available for patient privacy purposes, but are available from the corresponding authors upon reasonable request (R.J.G).

## Compliance with Ethical Standards

### Funding

This study was funded by U.S. Public Health Service research grant U01 CA143062 and R01 CA190105 (awarded to Dr. Gillies). This material includes work supported with resources and the use of facilities at the James A. Haley Veterans’ Hospital.

### Conflict of Interest

Robert J. Gillies declared a potential conflict with HealthMyne, Inc [Investor (major), Board of Advisors (uncompensated)]. Contents of this research do not represent the views of the Department of Veterans Affairs or the United States Government. The remaining authors declare no conflict of interest.

### Ethical approval

All procedures performed in studies involving human participants were in accordance with the ethical standards of the Institutional Review Board at the University of South Florida (USF) and the James A. Haley Veterans Hospital, and with the 1964 Helsinki declaration and its later amendments or comparable ethical standards.

### Informed consent

The requirement for informed consent was waived.

## Notes

Disclosure: This paper is supported by U.S. Public Health Service research grant U01 CA143062 and R01 CA190105 (principal investigator Dr. Gillies). Robert James Gillies declared a potential conflict with HealthMyne, Inc [Investor, Board of Advisors]. The remaining authors declare no conflict of interest.

