## Supplemental Methods, Tables, and Figures for "Radiomic and clinical predictors of cachexia in non-small cell lung cancer patients treated with immunotherapy"

.

### Supplementary Appendix E1

#### *Section 1: PET/CT imaging*

For the HLM patients, PET/CT images of the required quality (slice thickness ≤5 mm, no artifacts) from 9 different PET/CT equipment with 19 different scanners and reconstruction parameters, were collected to develop a more robust model. The 9 different PET/CT equipment include GEMINI TF TOF 16 [Philips], Discovery-600 [GE Medical Systems], Discovery ST [GE Medical Systems], Discovery LS [GE Medical Systems], Discovery STE [GE Medical Systems],CPS 1023 [CPS], BioGraph HiRes Model 1080 [Siemens], BioGraph TruePoint Model 1093 [Siemens], and Biograph40 mCT [Siemens]. All patients were fasted for at least 6h, and injected with 214–673 MBq of 18F-FDG depending on body weight at 93.46±24.86 min before data acquisition. The in-plane image resolution of PET and CT scans ranged from 2mm to 5mm, while the axial image resolution ranged from 2.73mm to 5.47 mm and 0.98mm to 5.47mm for PET and CT images, respectively.

For the VA patients, PET/CT images were acquired from Siemens Biograph Sensation 16 PET/CT. All patients were fasted for at least 6h, and injected with 342–598 MBq of 18F-FDG depending on body weight at 93.15±18.34 min before data acquisition. The in-plane image resolution of PET and CT scans were 2mm, while the axial image resolution was 2 mm and 0.98 mm for PET and CT images.

#### *Section 2: Image feature extraction*

Group1: Three dimensional PET imaging features. A total of 364 3D imaging features of the tumor from PET images, including 209 features (including 62 textural features, 48 statistical features, 42 morphological features, and 57 diagnostic features) described in ref (S1), and an additional 125 Laws features and 30 wavelet features (Section 3) were calculated.

Group II: Three dimensional CT imaging features. Similar to PET imaging features, the same set of 364 3D imaging features of the tumor from CT images was also extracted.

Group III: Three dimensional fusion imaging features. Fusion imaging plays an important role in clinic, since it can provide the location and boundary information from CT images in addition to the metabolic information from PET images. This is currently processed mentally by the nuclear medicine physician or oncologist. To perform quantitative fusion, we constructed fusion images based on $I_{fuse} = I_{PETnorm} + 0.6 \times I_{CTnorm}$, wherein $I_{PETnorm}$ and $I_{CTnorm}$ are normalized PET and CT pixel-wise image data[29]. From this fusion image, 62 textural and 3 statistical features (mean, minimum, and maximum) of the 364 features were calculated.

Group IV. Three dimensional habitat imaging features. Given that tumors are anatomically heterogeneous as a result of spatially variant cellular proliferation, necrosis, fibrosis, angiogenesis, molecular characteristics, and metabolism [S1], we divided the PET images using Otsu thresholding[30] into high and low metabolic (SUV) regions, as illustrated in Figure 2C. These sub-regions represent distinct habitats, which might respond differentially to therapy or drive progression [S2]. These two habitats were then mapped to the co-registered CT images, and two CT sub-images combined with the PET information were obtained. The CT sub-images corresponding to the high and low SUV regions in PET images were denoted CThigh and CTlow, respectively. Compared to the original CT images, these CT sub-images provide more information since they incorporated the metabolic information provided by PET images. Complementarily, the two new PET sub-images were also used and named PEThigh and PETlow. There were 62 textural features and 3 statistical features (mean, minimum, and maximum) extracted from each of these 4 sub-images.

Totally, 1053 features were generated, and all the training features were scaled into the range [0 1] with unity-based normalization, and the test features were normalized with the minimum and maxim values of training datasets in a similar way.

*S1. Alex Zwanenburg SL, Martin Vallières, Steffen Löck. Image biomarker standardisation initiative. arXiv:161207003 2017.*

*S2. Chicklore, S., et al., Quantifying tumour heterogeneity in 18F-FDG PET/CT imaging by texture analysis. Eur J Nucl Med Mol Imaging, 2013. 40(1): p. 133-40.*

*S3. Gatenby, R.A., O. Grove, and R.J. Gillies, Quantitative imaging in cancer evolution and ecology. Radiology, 2013. 269(1): p. 8-14.*

### *Section 3: Laws and wavelet features*

**Laws features:**

Laws features are constructed from a set of five one-dimensional filters, each designed to reflect to a different type of structure in the image. These one-dimensional filters are defined as E5 (edges), S5 (spots), R5 (ripples), W5 (waves), and L5 (low pass, or average gray value). By combining any three of the above 1-D convolution filters, we could obtain 125 3-D filters, and generated 125 filtered images. For each filtered image, the energy was calculated.

$$Energy = \frac{1}{M \times N \times L} \sum_{i}^{M} \sum_{j}^{N} \sum_{k}^{L} h^2(i, j, k) \quad (1)$$

Where M, N and L are filtered images dimensions, $h(i, j, k)$ means the filtered image.

**Wavelet features**

The discrete wavelet transform can iteratively decompose an image (3D) into four components. Each iteration splits the image both horizontally and vertically into low-frequency (low pass) and high frequency (high pass) components. Thus, four components are generated: a high-pass/high-pass component consisting of mostly diagonal structure, a high-pass/low-pass component consisting mostly of vertical structures, a low-pass/high-pass component consisting mostly of horizontal structure, and a low-pass/low-pass component that represents a blurred version of the original image. Subsequent iterations then repeat the decomposition on the low-pass/low-pass component from the previous iteration. For each filtered image, the energy was calculated.

$$Energy = \frac{1}{M \times N \times L} \sum_{i}^{M} \sum_{j}^{N} \sum_{k}^{L} h^2(i, j, k) \qquad (2)$$

Where M, N and L are filtered images dimensions, $h(i, j, k)$ means the filtered image.

### *Section 4: Radiomics signature calculation formulas*

The formula of RS is

$$RS = -0.70 \times PET_{LD} - 0.15 \times PET_{SRHGE} - 0.64 \times CT_{Energy} + 0.69 \times CT_{ZSN} - 0.30 \times CT_{max} - 0.30 \times PEThigh_{DD} + 0.35 \times PETlow_{LZLGE} + 0.018 \times CThigh_{mean} + 0.12 \times FUSE_{MHS}$$

Note. The feature is named as f_type, where f represents the feature name and type represents the type of the image (PET, CT, Fuse, PEThigh, PETlow, CThigh or CTlow images) used to calculate f.

LD means longest diameter; SRHGE means short run high grey level emphasis caculated from Grey-Level Run Length Matri; ZSN means Zone size non-uniformity Zone size non-uniformity emphasis caculated from gray level size zone matrix; DD means Degree of Direction caculated from Texture spectrum matrix; LZLGE means Large zone low grey level emphasis caculated from gray level size zone matrix; MHS means Micro-Horizontal Structrure calculated from Texture spectrum matrix.

### Supplementary Tables

Table S1. Demographic and clinical characteristics of VA IO-treated patients

| Characteristic | All(N=35) | Radiomics score | | *P* |
|---|---|---|---|---|
| | | High (N=18) | Low (N=17) | |
| Age(y) | | | | 0.91 |
| Mean ± SD | 71.40±7.19 | 71.00±6.70 | 71.82±7.87 | |
| Sex, NO. (%) | | | | NaN |
| Male | 35 (100) | 18 (100) | 17 (100) | |
| Female | 0 | 0 | 0 | |
| TNM stage | | | | 0.26 |
| III | 10 (28.57) | 7 (38.89) | 3 (17.65) | |
| IV | 25 (71.43) | 11 (61.11) | 14 (82.35) | |
| Histology (baseline), NO. (%) | | | | 0.018* |
| ADC | 19 (54.29) | 6 (33.33) | 13 (76.47) | |
| SCC | 16 (45.71) | 12 (66.67) | 4 (23.53) | |
| Smoke, NO. (%) | | | | 1.00 |
| Never | 1 (2.86) | 1 (5.56) | 0 | |
| Former | 34 (97.14) | 17 (89.47) | 17 (100) | |
| ECOG PS, NO. (%) | | | | 0.024* |
| 0 | 7 (20.00) | 2 (11.11) | 5 (29.41) | |
| 1 | 22 (62.86) | 10 (55.56) | 12 (70.59) | |
| >=2 | 6 (17.14) | 6 (33.33) | 0 | |
| Weight change within 6 months | | | | 0.067 |
| Mean ± SD | -1.95±8.25 | -4.56±8.00 | 0.65±7.91 | |
| Cachexia, NO. (%) | | | | 0.027* |
| Yes | 14 (46.67) | 10 (55.56) | 4 (23.53) | |
| No | 16 (53.33) | 5 (27.78) | 11 (64.71) | |
| Unknown | 5 (16.67) | 3 (16.67) | 2 (11.76) | |
| Progression-free Survival | | | | 0.002* |
| Median (IQR) | 8.13(2.87, 13.4) | 4.77 (2.50, 8.13) | 12.55 (9, NR) | |
| Overall Survival | | | | 0.039* |
| median (95%CI) | 13.10 (6.63, NR) | 8.37(5.63, 17.00) | 22.17 (13.10, NR) | |
| RS | | | | <.001* |
| Median (IQR) | 0.41(0.35,0.47) | 0.46 (0.43,0.52) | 0.21 (0.35,0.38) | |

Note. * means P value <.05. The comparison of age, RS and Weight change between two groups was performed with two-sided Wikcoxon sign rank test, and the rest variables were compared with two-sided Fisher’s test. IQR is short for interquartile range. ADC is short for adenocarcinoma and SCC is short for squamous cell carcinoma. NA means not available. NR means not reached.

Table S2. Logistic regression analysis of risk factors for cachexia prediction

| | Univariable | | Clinical Multivariable | | Radiomics Multivariable | |
|---|---|---|---|---|---|---|
| | Odds Ratio (95% CI) | p | Odds Ratio (95% CI) | p | Odds Ratio (95% CI) | p |
| RS | 7.23 (3.18~16.43) | <.001* | | | 5.49 (2.30~13.08) | <.001 |
| SUVmax | 0.95 (0.87~1.05) | 0.34 | | | | |
| BMI category | 0.59 (0.38~0.91) | 0.018* | 0.63 (0.39~0.99) | 0.049 | 0.65 (0.39~1.06) | 0.086 |
| SMI category | 0.67 (0.32~1.38) | 0.28 | | | | |
| Sex | 0.91 (0.44~1.89) | 0.80 | | | | |
| Age | 0.98 (0.94~1.02) | 0.34 | | | | |
| Smoke | 1.20 (0.62~2.33) | 0.60 | | | | |
| COPD | 2.18 (0.83~5.74) | 0.12 | | | | |
| ECOG PS | 2.89 (1.21~6.89) | 0.017* | 2.35 (0.94~5.87) | 0.066 | 1.59 (0.59~4.24) | 0.36 |
| Distant Metastasis | 1.54 (1.09~2.18) | 0.015* | 1.50 (1.05~2.15) | 0.025 | 1.31 (0.89~1.92) | 0.18 |
| Histology | 2.14 (0.99~4.60) | 0.052 | | | | |
| MTV | 1.01 (0.998-1.01) | 0.13 | | | | |
| Volume | 1.00 (0.999-1.01) | 0.16 | | | | |
| Constant | | | 0.40 | 0.29 | 0.29 | 0.19 |

* P value <.05
